# Research on the Influence of Information Diffusion on the Transmission of the Novel Coronavirus (COVID-19)

**DOI:** 10.1101/2020.03.31.20048439

**Authors:** Shanlang Lin, Chao Ma, Ruofei Lin, Junpei Huang, Ruohan Xu, Aini Yuan

## Abstract

With the rapid development of mobile Internet in China, the information of the epidemic is full-time and holographic, and the role of information diffusion in epidemic control is increasingly prominent. At the same time, the publicity of all kinds of big data also provides the possibility to explore the impact of media information diffusion on disease transmission. This paper explores the mechanism of the influence of information diffusion on the spread of the novel coronavirus, develops a model of the interaction between information diffusion and disease transmission based on the SIR model, and empirically tests the role and mechanism of information diffusion in the spread of COCID-19 by using econometric method. The result shows that there was a significant negative correlation between the information diffusion and the spread of the novel coronavirus; The result of robust test shows that the spread of both epidemic information and protection information hindered the further spread of the epidemic.

## 1. Introduction

Since December 2019, a number of cases of viral pneumonia with unknown causes have been found in Wuhan, which has been confirmed as the novel coronavirus 2019 (hereinafter referred to as COVID-19). Despite strict interventions such as isolation treatment and traffic control, the epidemic spread rapidly to all provinces and cities in the country at an unprecedented rate. As of February 25, 2020, according to the reports of 31 provinces (autonomous regions, Municipality) and Xinjiang Production and Construction Corps, a total of 77271 confirmed cases, 3434 suspected cases, 2596 death, and 25065 cured cases have been reported. This is another major public health emergency in China after the attack of the SARS virus in 2003.

Since the outbreak of COVID-19 occurred around the Spring Festival, the scale of population movement was large and the frequency was high, which also increased the difficulty of epidemic prevention and control. In order to prevent the spread of the epidemic, the central government has taken unprecedented prevention and control measures, including setting up designated admission hospitals, expanding the supply of beds in the hospital, coordinating the dispatch of medical prevention and control materials, extending the Spring Festival holiday, implementing peak staggering return, measuring the temperature of vehicles and stations, disinfecting, ventilating, etc. The National Health Commission has also sent a number of supervision teams to hospitals and disease control agencies to conduct on-site supervision. Wuhan also announced the closure of the city on January 23, suspending urban public transport and strictly controlling the access of people inside and outside Wuhan. Subsequently, 31 provinces, regions and cities in the country successively launched the level 1 emergency response to public health emergencies, strictly controlled the transmission of the virus, and made every effort to prevent the further spread of the epidemic.

At the early stage of the epidemic (December 31 to January 20), due to the lack of public reports in Wuhan’s official media, the information of the epidemic was mainly disclosed by the media, so the epidemic was not paid attention by the public, which delayed the best time to expand the social impact, thus leading to the further spread of the epidemic. After January 20, with the outbreak of the epidemic and the release of information, COVID-19 epidemic information became the most concerned information of the public, and media reports entered a white-hot stage. The emergence of the novel coronavirus pneumonia has also caused widespread panic among the public. The official media, micro-blog, WeChat and other media have followed up the reports of real-time epidemic, new symptoms and prevention measures, and timely conveyed clear and positive information to the society and advised the public to protect themselves and to view the epidemic more objectively and objectively. Will the information diffusion help to eliminate rumors and guide the public to do a good job in protection and further inhibit the spread of the epidemic? Therefore, understanding the impact of information diffusion on epidemic transmission can help improve the prediction of epidemics and find preventive measures to slow down the spread of diseases.

Therefore, this paper studies the problems above. The innovation points of the research are as follows: First, combing the epidemic theory, complex network analysis and the temporal and spatial background of the COVID-19 spread, we comb the mechanism of epidemic transmission; Second, we use econometric method to conduct a regress, and get the basic conclusion that information diffusion can effectively reduce the spread of COVID-19; third, using big data mining technology, Baidu search index, Baidu population migration, prevention and control data during the epidemic were mined through Baidu and government information websites at all levels.

The remaining sections of the paper are as follows: the second section combs the research literature and puts forward the hypothesis; the third section is the research design, which describes the model construction, data source, main variable calculation, and statistical description; the fourth section is empirical analysis. The fifth section is a conclusion and discussion.

## 2. Literature Review and Hypotheses

### 2.1 Literature Review

Since the beginning of the 21st century, due to the outbreak of SARS, avian influenza, novel H1N1 influenza and Ebola cross the world, the public has been increasingly concerned about the emerging infectious diseases, and the problem of disease transmission has been widely studied (Mao & Yang, 2012). In general, the spread of an epidemic is considered to be a dynamic process in which the disease passes from one individual to another through contact between individuals on the contact network (kleczkowski et al., 2011). Disease transmission often occurs in a dynamic social environment, and individual health behavior decision-making is guided by cultural norms, peer behavior and media reports (Kim et al., 2019). Although vaccination is a major strategy to protect individuals from infection, the development, testing and production of new vaccines often take a long time (stohr & esveld, 2004). Before getting enough vaccines, the best protection for individuals is to take preventive actions, such as wearing masks, washing hands frequently, taking drugs, avoiding contact with patients, etc. (Centers for Disease Control and prevention, 2008). The historical experience of SARS tells us that effective national control measures, such as early identification and isolation of SARS cases, tracking and isolation of the contacts, screening of travelers, and raising public awareness of risk, can help to contain the spread of the virus (Ahmad et al., 2009).

As the public gradually realized the importance of personal behavior in preventing the spread of infection, researchers began to explore the mathematical model of disease transmission including personal behavior. These models have been used to guide strategies for disease transmission control (vardavas et al., 2007) and quantify the role of individual protective measures in controlling several outbreaks, including the Ebola virus outbreak in West Africa in 2014 (fast et al., 2007), the SARS outbreak in Hong Kong in 2003 (Riley, 2003) and the H1N1 outbreak in central Mexico in 2009 (springborn et al., 2007). Saunders et al. (2017) also tested the effectiveness of personal protective measures in preventing the spread of pandemic influenza in humans. Recently, the research on the outbreak of COVID-19 from the perspective of transmission dynamics is also increasing. Sun et al. (2020) evaluated the epidemiological trend of COVID-19 based on the public epidemic data, and studied the outbreak progress in all parts of China.

Understanding the impact of the media on the spread of the disease can help improve the prediction of epidemics and identify preventive measures to slow the spread of the disease. Many models also link the disease-related media transmission with the protection function, usually assuming that the influence of media will reduce the effective transmission rate and slow down the spread of diseases. These studies indicate that the impact of media increases with the number of people infected (Sun et al., 2011; Liu et al., 2007; Cui et al., 2008), or both with the number and the rate of change (tchuenche & bauch, 2012; Xiao et al., 2015). When the number of cases is high or the prevalence of diseases increases rapidly, the information diffusion slows down the spread of diseases and creates interesting disease transmission dynamics, such as multi-wave outbreaks (Liu et al., 2007; Cui et al., 2008). However, it is not clear whether the media function formalization proposed by the model fully reflects the actual influence. The choice of media function directly affects the form of disease transmission, making the accurate parameterization of the media is the key (collison et al., 2014).

However, most of the current researches only focus on the development of the disease itself on the complex network, as well as the impact of protective measures on the spread of the disease. There are relatively few studies on the spread of disease-related information, and only a few of them are carried out through numerical simulation with the preconditions which are too idealized and too dependent on the setting of parameters. In addition, the model has just begun to consider how to combine the data from actual media reports (collison et al., 2015), lacking the econometric analysis based on real-time data. In decade years, China’s Internet has experienced unprecedented development, various online social media based on the Internet (such as major search engines, social networking sites, news sites, etc.) have been integrated into people’s daily life, providing a broad platform for the dissemination of various information. Compared with the outbreak of SARS in 2003, the economic link between regions are increasingly close, and during the Spring Festival in China, the population flow is more frequent. Although the government has taken unprecedented measures to prevent and control the epidemic, and the official media and social media have timely released the latest epidemic information, the actual effect needs to be further tested. By combing and integrating the existing literature, we propose the hypothesis that information diffusion is helpful to curb the spread of new coronavirus.

### 2.2 Hypothesis

As shown in the figure 2, there is a clear correlation between the COVID-19-related information and the spread of the epidemic. This indicates that it is feasible to study the development of the epidemic through the Network public opinion information. Further, we visualized the geographic distribution of the epidemic and information (as shown in figure 3). Overall, there is a correlation between the two. It can be seen that the place with the most public opinion information is actually not the place where the epidemic is the most serious (such as Hubei Province, but it does not rule out that the epidemic area has no time to process the information), but the first-tier provinces such as Beijing, Shanghai and Guangdong (The situation in these cities is still severe, but it is relatively stable compared to Hubei Province). Therefore, this article proposes a hypothesis: the spread of epidemic information will inhibit the spread of the epidemic.

**Figure 1.**
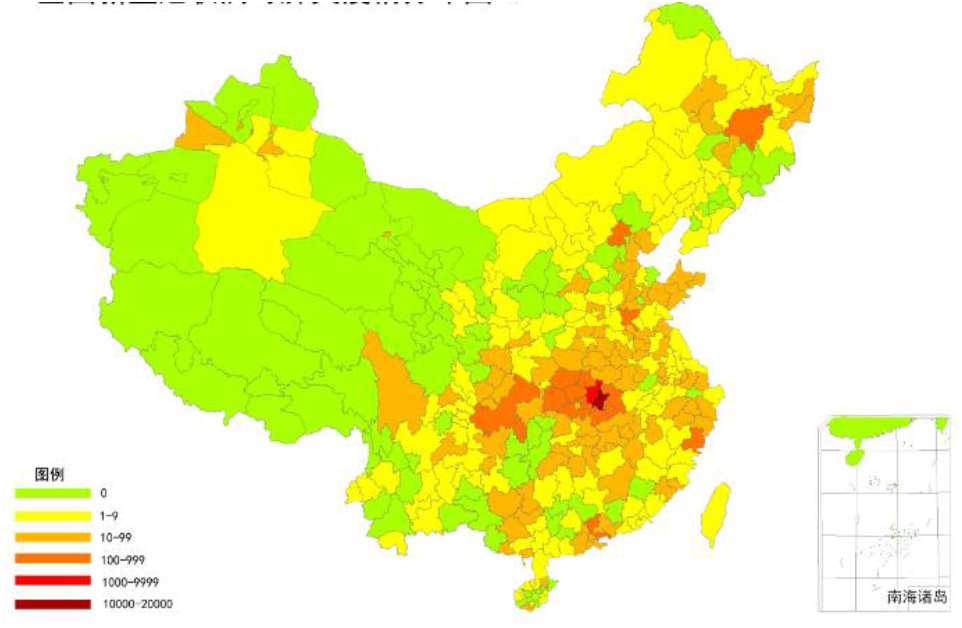
Distribution of COVID-19 Outbreaks (as of 25 February)

**Figure 2.**
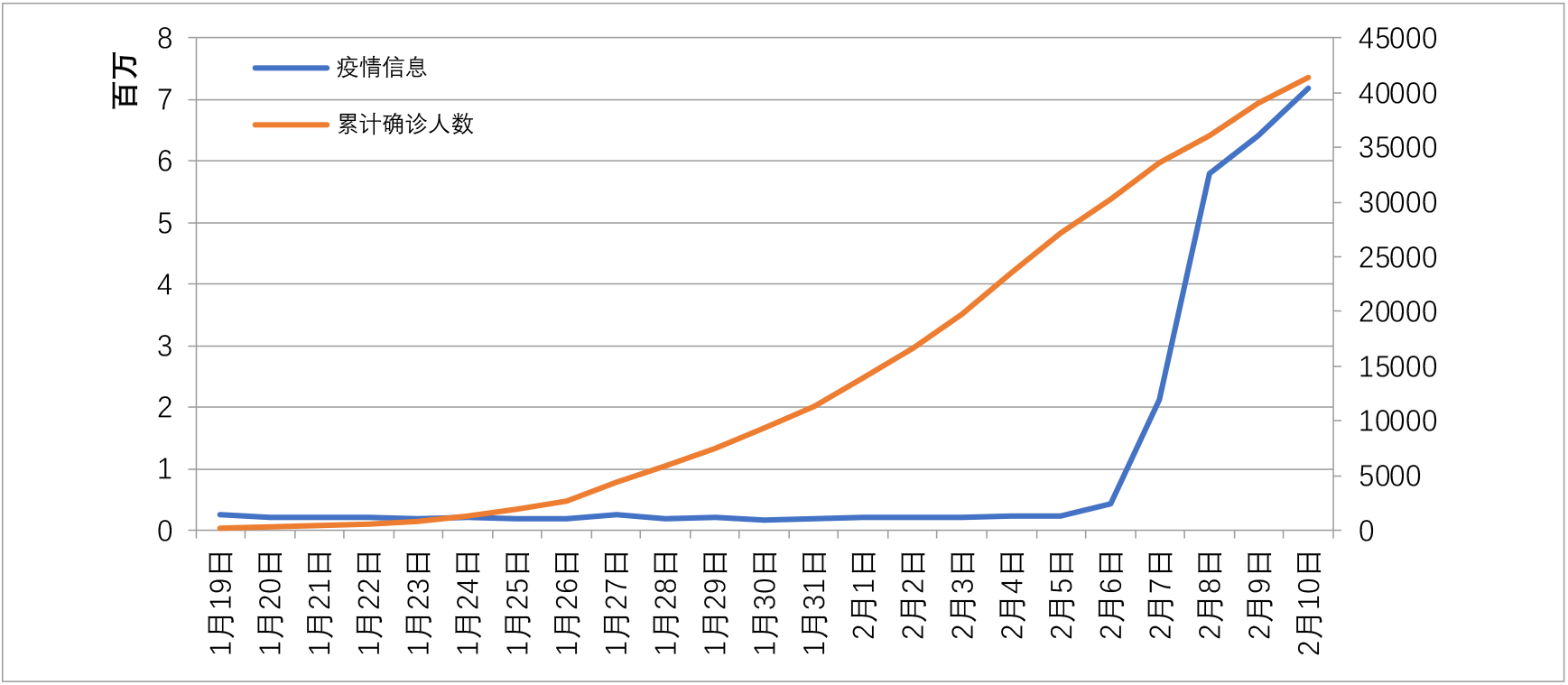
Confirmed Cases and the Public Opinion Information in China

**Figure 3.**
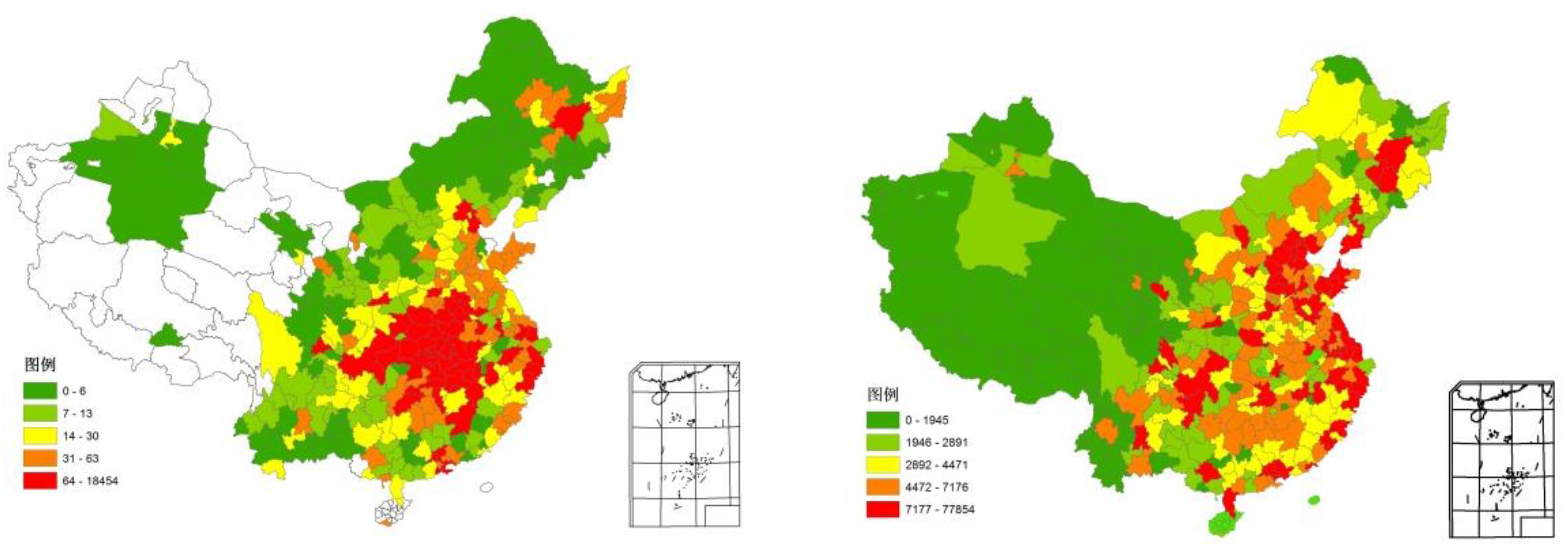
Geographical Distribution of the Number of Confirmed Cases and Public Opinion Information in China (February 10)

### 2.3 Analysis of Mechanism

The mechanism of the influence of information dissemination on the spread of COVID-19 is as follows:

When a disease spreads among people, information about the disease spreads immediately. In the early days of the outbreak, a small amount of information spread, mainly about the popular science of the virus and the current infection dynamics. The information has improved the public’s awareness of the virus. Many people choose to continue to pay attention to the epidemic-related information, and respond faster when the outbreak later occurs, reducing the probability of infection.

In the stage of the outbreak, information about it will also be spread on major social media. When individuals learn about the existence of the disease, they will change their behavior, such as wearing a mask and vaccinating to avoid being infected, which can have an impact on the spread of the disease. The susceptible (or infected person) who knows the information of the epidemic will break the connection with the around infected person (or susceptible person) in order to prevent the further spread of the disease.

After receiving the epidemic-related information, governments at all levels also quickly launched emergency plans, and gradually adopted a series of measures such as early identification and isolation, traveler screening, closing of public places, and even “lockdown” the cities in accordance with the deterioration of the epidemic to limit traffic and population flow, greatly reducing further transmission of the epidemic.

## 3. Methodology

### 3.1 Model Construction

In the current literature, models of disease transmission and behavioral spread have been developed for decades, all are based on human networks ((Deffuant, Huet & Amblard, 2005; Keeling & Eames, 2005). However, few people committed to combining information diffusion and human behavior, considering these two interactive processes. In fact, when a disease-related information spreads among people, people will naturally take some precautionary measures to counteract it, which in turn limits the spread of the disease.

In order to study the impact of information diffusion on epidemic outbreaks, this paper draws on the conceptual framework of Mao & Yang (2012) to develop a model of the interaction between disease and information. We divided the population into two categories according to their health status: the susceptible (S) and the (I); according to the level of information obtained, they were divided into two categories: conscious (+) and unconscious (-). Therefore, the population in the entire society can be divided into four states: (1) S−: unconscious susceptible; (2) S +: conscious susceptible; (3) I−: unconscious infected; (4) I +: conscious infected. As shown in the figure 5, the transmission process of COVID-19 including the information diffusion is proposed. The entire propagation process of the model is described as follows:

**Figure 4.**
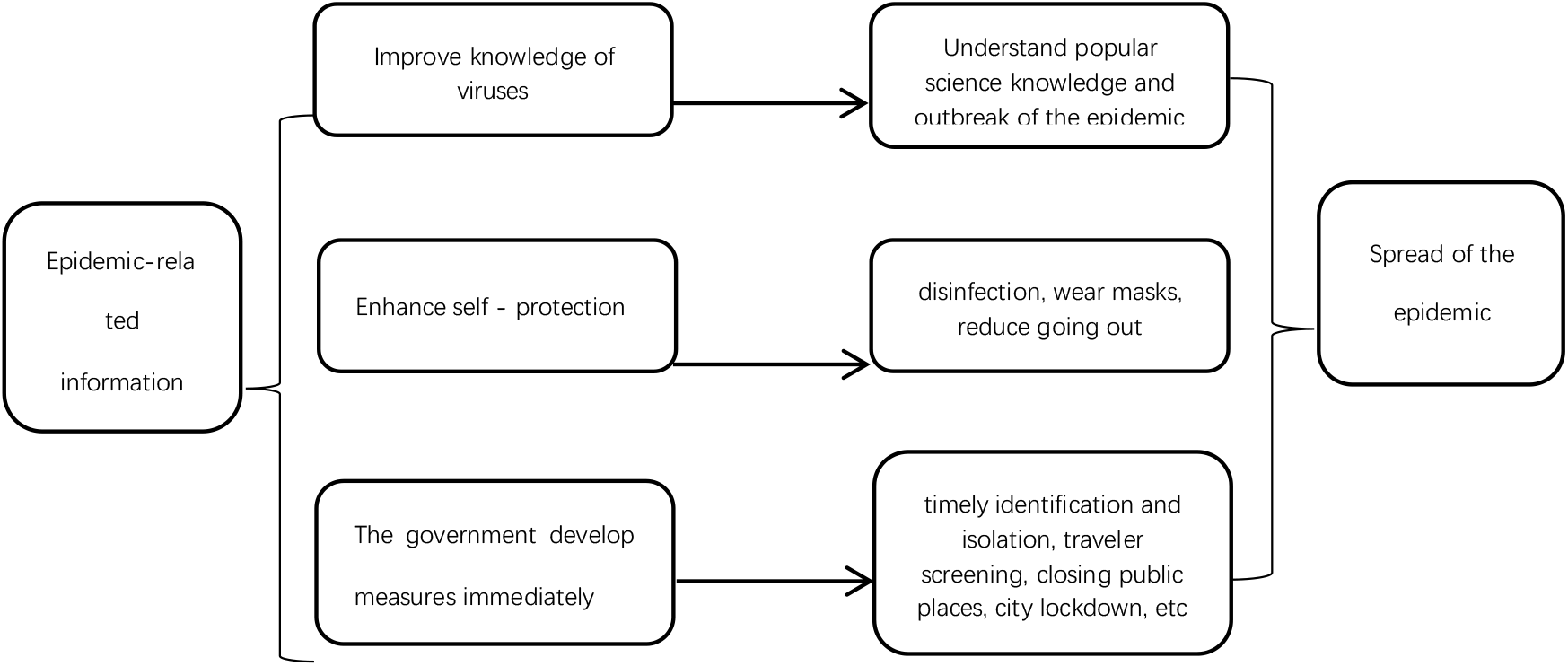
The Transmission mechanism

**Figure 5.**
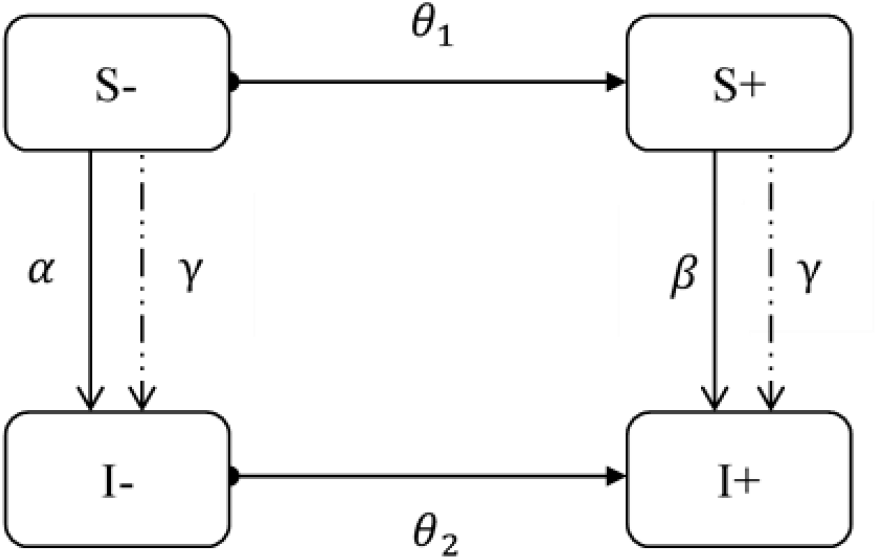
Schematic Diagram of COVID-19 Spread and Information Diffusion

Information diffusion: In this process, the Internet and other media reported epidemic-related information on a large scale. Some unconscious individuals in the susceptible (*θ*_1_) received the information and became conscious individuals; and some of the unconscious individuals in the infected (*θ*_2_) received the information and become conscious individual.

Spread of the COVID: In this process, unconsciously susceptible people (S-) are infected with the virus with probability *α*, while consciously susceptible people usually take self-protection measures such as wearing a mask, reducing going out, disinfection, etc. so they are infected with the probability of *β* (*β* < *α*); due to isolation treatment for the infected patients, they (I− and I +) pass the disease to surrounding susceptible (S + and S−) with the same probability of transmission *γ*. Since the reinfection process after recovery of patients with COVID-19 is not clear, here we don’t take it into consideration.

Therefore, the information diffusion can reduce the spread of the epidemic in two ways. First, information diffusion will cause some conscious and susceptible people (*β* •*S* +) to take proactive protective measures to prevent infection; second, information diffusion will change some unconsciously susceptible people (*θ*_1_ •*S −*) into consciously susceptible people, and then take protective measures to reduce some (*θ*_1_; •*S −* •*β*) infection.

According to the hypothesis above, the empirical model of the influence of information diffusion on the spread of COVID-19 is as follow:

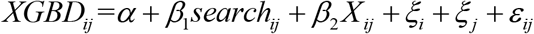

Where, *i* denotes date, *j* denotes city; *XGBD* is the spread of COVID-19, which is measured by the number of cumulative and newly confirmed cases published by the National Health Commission each day; *X* is control variables, including traffic control (*traf* _ *con*), self-control (*self* _ *seg*), movement of population (*migration*), Population inflow rate of Wuhan (*ratio*), and GDP per capita (*pergdp*). *ξ*_*I*_ is time fixed effect, *ξ* _*j*_ is city fixed time, *ε*_*ij*_ the random error term.

### 3.2 Data Resource

In order to quantitatively explore the relationship between the spread of the COVID-19 and information diffusion, we first visited the Baidu Index website through Python to obtain the Baidu search index2 of the keywords related to the epidemic during the outbreak from January 19 to February 10, 2020 to measure the level of information diffusion. The data of infected case during the corresponding period mainly come from the daily epidemic data released by the National Health Commission. The self-control and traffic control data come from the public information of each city’s Health Commission website and government website on taking preventive and control measures, and they are scored uniformly according to the degree of control, and the corresponding values are added up. The national migration data and Wuhan’s outflow data come from Baidu Migration. The control variables at the city level come from China City Statistical Yearbook. In addition, cities without outbreaks were also excluded. After collation, 6417 observations from 301 cities were finally obtained.

### 3.3 variable description and measures

Coronavirus transmission (*XGBD*): The number of the infected in this article is used to indicate the transmission of the virus. After the outbreak of the epidemic, the National Health Commission provided daily outbreak data. Therefore, we have compiled a list of the daily number of cumulative and newly confirmed in prefecture-level cities from January 19 to February 10, 2020.

Information diffusion (*search*): The main explanatory variable---information diffusion in this paper is measured by the number of searches for epidemic-related information by the national people every day during the epidemic. Different from the media data (***) used in previous studies as the level of information diffusion, the search index can better reflect people’s acceptance of information diffusion. Therefore, based on the search services provided by Baidu Index, six epidemic-related terms of “the novel coronavirus”, “pneumonia”, “Zhong Nanshan”, “pneumonia symptoms”, “masks” and “correct wearing of masks” were selected as search terms, and the search index during the epidemic period (January 19-February 10) was crawled by Python, and the daily level of information diffusion of prefecture-level cities was summed up. In addition, the search index is divided into two categories, one is about the information on epidemic with “the novel coronavirus, pneumonia, Zhong Nanshan” (*search*1), and the other is about the protection with “pneumonia symptoms, correct wearing of masks and masks” (*search*2).

Figure 5 Distribution of Search Index of prefecture-level administrative units in China (February 10)

Traffic Control (*traf* _ *con*) and Self-control (*self* _ *seg*): This paper collects, summarizes and collates the epidemic prevention and control mechanisms published by the Emergency Command of the Novel Coronavirus Pneumonia Prevention and Control in each province and city, mainly including traffic control and self-control. According to these preventive and control measures taken by all prefectural administrative regions in the country, they are classified and scored into 15 items (see Table 1), each with a score of 1, starting from the time when each measure is implemented until the measure is cancelled. For example, Shanghai began to implement the “isolation of close contacts of confirmed patients” for 14 days on January 21. Since this measure belongs to “social isolation”, the “social isolation” score of Shanghai was 1 from January 21. On January 24, Shanghai began to implement the “partial cessation of public places in the city”, then the “social isolation” was added 1 point from January 24, and so on. Finally, traffic control was carried out separately. The scores of each measure of social alienation were summed up.

**Table 1.**
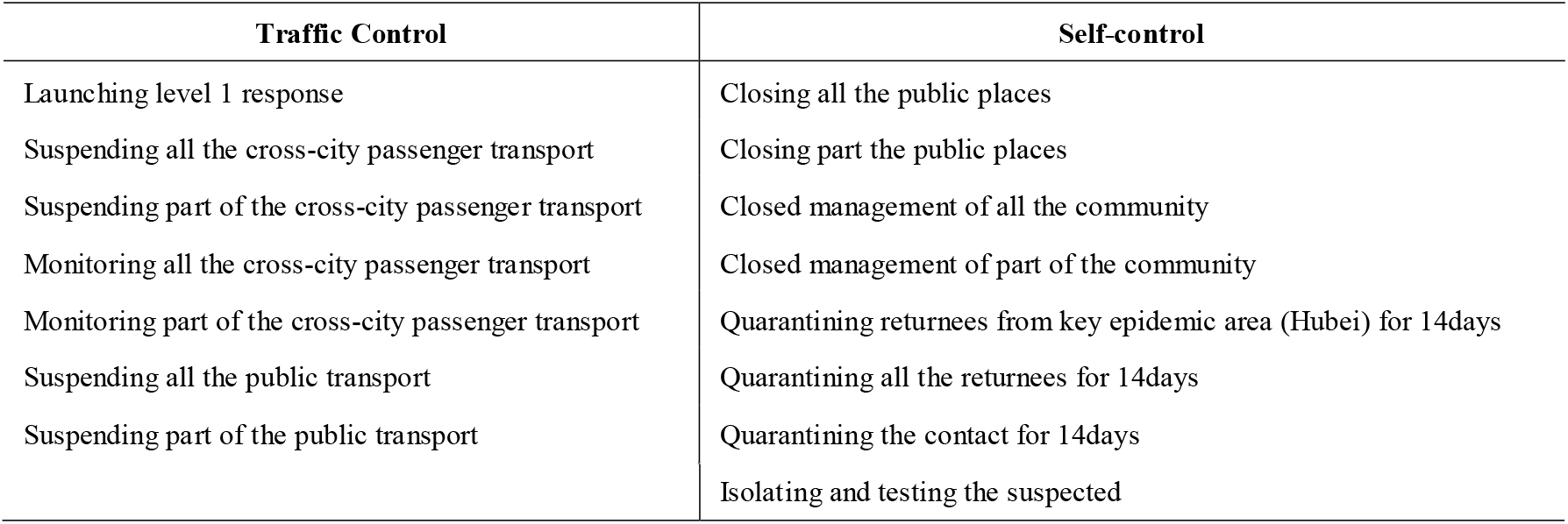
Items of Traffic Control and Self-control

Population Flow (*migration*): As the epidemic occurred during the Spring Festival Movement in China, the large-scale population flow provided favorable conditions for the spread of the virus, and reasonable control of population flow helped to slow down the spread of the epidemic. Baidu Migration Big Data provides a migration index that reflects the scale of population migration into or out, and is comparable between cities. Therefore, the migration indexes of population moving in and out of prefecture-level cities were obtained respectively during the epidemic respectively and the indicators reflecting the overall population flow status of the city were summed up.

Other control variables. (1) The influx of population in Wuhan (*ratio*). Wuhan is the city of the outbreak, and the influx of Wuhan population may lead to the cross-city transmission of the epidemic. The big data of Baidu Migration provides the destination and proportion of population migration in Wuhan every day. This paper selects the proportion of population flow in Wuhan to other cities to represent the population inflow in Wuhan; (2) GDP per capita (*pergdp*). GDP per capita reflects the level of urban social and economic development, while cities with high economic development tend to have more complete epidemic prevention facilities and stronger epidemic prevention capacity.

### 3.4 statistical description

## 4. Empirical analysis

### 4.1 Benchmark regression

According to the econometric model constructed above, benchmark regression was performed by controlling time and urban fixed effects separately. Column (1) in Table 3 performed a direct regression on the spread of information and the spread of the COVID-19, and the results were statistically significant and negative. Column (2) showed results when added traffic control and personal control variables. Column (3) showed the results after adding further variables such as population flow, Wuhan inflow, and GDP per capita based on column (2). The regression results are also statistically significant and negative. This indicates that after controlling other factors affecting the COVID-19, the spread of information has significantly reduced the spread of COVID-19 in the country. The regression coefficients of traffic control and personal control were significantly negative, indicating that after implementing a first-level response measures such as urban traffic control, segregated observation, and closed communities adopted by local governments were significantly effective, reducing the spread of the COVID-19. In terms of variables reflecting population migration, the coefficient of urban population migration variable is significantly negative, indicating that the decrease in population inflows and outflows has also reduced the spread of COVID-19; the coefficient of Wuhan population inflow variables is significantly positive, indicating that the population inflow in Wuhan has accelerated the spread of the virus to a certain extent.

**Table 2.** Statistical Description of Variables

| Variable |  | Obs | Mean | Std.Dev. | Min | Max |
| --- | --- | --- | --- | --- | --- | --- |
| qzrs | Cumulative number of confirmed cases | 6417 | 0.351 | 2.479 | 0 | 66.63 |
| xzqz | Newly number of confirmed cases | 6417 | 0.0540 | 0.474 | 0 | 28.63 |
| search | Information Diffusion | 6417 | 56.77 | 38.20 | 1.017 | 317.7 |
| search1 | Information Diffusion-epidemic | 6417 | 49.65 | 34.18 | 0.814 | 294.2 |
| search2 | Information Diffusion-prevention | 6417 | 7.112 | 4.665 | 0 | 54.57 |
| sea_pergdp | Interaction term of Search Index | 6417 | 344.4 | 362.7 | 3.122 | 3864 |
| sea1_pergdp | Interaction term of Search Index | 6417 | 303.0 | 327.1 | 2.535 | 3554 |
| sea2_pergdp | Interaction term of Search Index | 6417 | 41.41 | 38.81 | 0 | 436.7 |
| migration | population flow rate | 6417 | 1.720 | 2.618 | 0.0272 | 31.23 |
| ratio | Population inflow rate of wuhan | 6417 | 0.307 | 1.737 | 0 | 23.86 |
| traf_con | Traffic Control | 6417 | 2.418 | 1.633 | 0 | 4 |
| self_seg | Self-control | 6417 | 2.478 | 2.061 | 0 | 6 |
| pergdp | GDP per capita | 6417 | 5.779 | 3.180 | 1.520 | 18.31 |

**Table 3.** Benchmark regression results

|  | (1)<br>qzrs | (2)<br>qzrs | (3)<br>qzrs | (4)<br>qzrs |
| --- | --- | --- | --- | --- |
| search | -0.0104**<br>(-2.26) | -0.00988**<br>(-2.20) | -0.00659***<br>(-3.48) | -0.00603***<br>(-3.21) |
| traf_con |  | -0.142***<br>(-2.78) | -0.0860**<br>(-2.27) | -0.0727**<br>(-1.97) |
| self_seg |  | -0.113***<br>(-4.72) | -0.0796***<br>(-4.21) | -0.0592***<br>(-3.32) |
| migration |  |  | -0.0458***<br>(-3.79) | -0.0271***<br>(-3.05) |
| ratio |  |  | 3.026***<br>(7.65) | 3.027***<br>(7.62) |
| pergdp |  |  | 0.0709*<br>(1.87) | 0.0151<br>(0.53) |
| _cons | 0.00462<br>(0.02) | 0.157<br>(0.70) | -1.035***<br>(-2.78) | -0.585*<br>(-1.89) |
| Fixed time | YES | YES | YES | YES |
| Fixed region | YES | YES | YES | YES |
| *N* | 6417 | 6417 | 6417 | 6394 |
| *R*<sup>2</sup> | 0.5266 | 0.5302 | 0.6787 | 0.6870 |

Wuhan is the place where the disease first spread on a large scale. The timing of the initial confirmed cases is unknown, and the lack of medical equipment and supplies after the outbreak has caused the number of confirmed cases to be lower than the actual number of confirmed cases. Therefore, in column (4), we exclude the data of Wuhan City, and the regression results are still stable.

### 4.2 Endogenous solutions and robustness tests

#### (1) Information classification robustness test

In order to learn more about the impact of information diffusion on the spread of COVID-19, this article divides the information represented by the Baidu search index into two categories. One is information on COVID-19 that the public desires to know (Search 1). The other is the self-protection information (search2) during the epidemic search by the public in order to prevent themselves from being infected by the virus.

Columns (1) and (2) in Table 4 are the results of robustness test which shows the impacts of the spread of epidemic information on the spread of COVID-19, the regression coefficients are significantly negative. Columns (3) and (4) are the results of robustness test which shows the impacts of the self-protection information diffusion on the spread of COVID-19, and the regression coefficients are also significantly negative. It shows that both the spread of virus information and self-protection information has hindered the further spread of the COVID-19.

**Table 4.** Regression results of different kinds of information t statistics in parentheses * p < 0.1, ** p < 0.05, *** p < 0.01

|  | (1) | (2) | (3) | (4) |
| --- | --- | --- | --- | --- |
|  | qzrs | qzrs | qzrs | qzrs |
| search1 | -0.00873<sup>*</sup><br>(-1.91) | -0.00567<sup>***</sup><br>(-2.90) |  |  |
| search2 |  |  | -0.184<sup>***</sup><br>(-3.40) | -0.112<sup>***</sup><br>(-5.12) |
| traf_con |  | -0.0871<sup>**</sup><br>(-2.29) |  | -0.0973<sup>***</sup><br>(-2.61) |
| self_seg |  | -0.0795<sup>***</sup><br>(-4.21) |  | -0.0832<sup>***</sup><br>(-4.37) |
| migration |  | -0.0479<sup>***</sup><br>(-3.94) |  | -0.0329<sup>***</sup><br>(-2.78) |
| ratio |  | 3.031<sup>***</sup><br>(7.64) |  | 2.957<sup>***</sup><br>(7.66) |
| pergdp |  | 0.0661<sup>*</sup><br>(1.74) |  | 0.0311<br>(0.89) |
| _cons | -0.0235<br>(-0.12) | -0.959<sup>***</sup><br>(-2.58) | -0.446<sup>***</sup><br>(-3.20) | -0.952<sup>***</sup><br>(-2.61) |
| Fixed time | YES | YES | YES | YES |
| Fixed region | YES | YES | YES | YES |
| N | 6417 | 6417 | 6417 | 6417 |
| R<sup>2</sup> | 0.5254 | 0.6783 | 0.5393 | 0.6831 |

#### (2) Robustness test of new cases

The data released by the National Health and Construction Commission includes the daily number of newly diagnosed patients, which can better reflect the spread of the epidemic every day. Therefore, we use the new confirmed number instead of the cumulative confirmed number in the previous model for further robustness testing. The regression results when use the comprehensive search index in column (2) in Table 5 are significantly negative, and the regression results when use the search index of disease information and protection information in column (3) and column (4) are also significantly negative, this further indicats that the regression results are robust and the spread of information can reduce the spread of the COVID-19.

**Table 5.** Regression results of new diagnoses t statistics in parentheses * p < 0.1, ** p < 0.05, *** p < 0.01

|  | (1)<br>xzqz | (2)<br>xzqz | (3)<br>xzqz | (4)<br>xzqz |
| --- | --- | --- | --- | --- |
| search_ | -0.00115*<br>(-1.72) | -0.000665**<br>(-1.98) |  |  |
| search1 |  |  | -0.000632*<br>(-1.76) |  |
| search2 |  |  |  | -0.00840**<br>(-2.55) |
| traf_con |  | -0.0108*<br>(-1.77) | -0.0108*<br>(-1.77) | -0.0119*<br>(-1.90) |
| self_seg |  | 0.00569<br>(1.49) | 0.00570<br>(1.49) | 0.00541<br>(1.43) |
| migration |  | -0.00754***<br>(-3.34) | -0.00769***<br>(-3.40) | -0.00678***<br>(-2.88) |
| ratio |  | 0.381***<br>(3.66) | 0.381***<br>(3.66) | 0.376***<br>(3.62) |
| pergdp |  | 0.00328<br>(0.52) | 0.000000326<br>(0.50) | -6.62e-08<br>(-0.10) |
| _cons | -0.0154<br>(-0.50) | -0.0563<br>(-0.96) | -0.0540<br>(-0.88) | -0.0388<br>(-0.61) |
| Fixed time | YES | YES | YES | YES |
| Fixed region | YES | YES | YES | YES |
| N | 6417 | 6417 | 6417 | 6417 |
| R<sup>2</sup> | 0.3934 | 0.4093 | 0.4092 | 0.4098 |

## 5. Conclusions and discussions

### 5.1 Conclusion

This paper draws on the model of behavioral dynamics, uses econometric methods and high-frequency data such as new coronavirus epidemic data published by the National Health and Medical Commission, Baidu search index and Baidu migration index to explore the relationship between information diffusion and the spread of COVID-19 Relationship. Studies show that: Firstly, after fixing time and cities and controlling other variables that affect the spread of New Coronavirus, the spread of information significantly reduces the spread of COVID-19. After excluding Wuhan from the sample, the regression results are still robust. Secondly, two robustness tests of information classification and new confirmed diagnoses show that both the spread of epidemic information and self-protection information have significantly reduced the further spread of the COVID-19. This shows that when the epidemic occurs, the timely and accurate spread of information plays an important role in the prevention and control of the epidemic.

### 5.2 Discussion

Excessive and inaccurate spread of epidemic information may also bring unexpected counter-effects, such as causing panic among the people, causing snatching of living materials, and curbing economic activities. Therefore, while information diffusion has played a role in the practice of epidemic prevention and control, it may be possible to take interventions to reduce the adverse effects. This article puts forward the following suggestions: (1) The government convenes a press conference in a timely manner to disclose the epidemic situation information and make the information transmission more transparent. The state-run media played a role of weathervane, they need to timely follow up the epidemic report to let public learn about the virus. (2) Do a good job of information management and control. Official authority, hospital and well-known experts need to timely deny a rumor for various purposes to avoid unhealthy social impact. (3) In view of the differences in urban and rural internet penetration rates, governments at all levels need to issue official documents timely to transfer information to rural areas.

## Data Availability

In order to quantitatively explore the relationship between the spread of the COVID-19 and information diffusion, we first visited the Baidu Index website through Python to obtain the Baidu search index of the keywords related to the epidemic during the outbreak from January 19 to February 10, 2020 to measure the level of information diffusion. The data of infected case during the corresponding period mainly come from the daily epidemic data released by the National Health Commission. The self-control and traffic control data come from the public information of each city’s Health Commission website and government website on taking preventive and control measures, and they are scored uniformly according to the degree of control, and the corresponding values are added up. The national migration data and Wuhan's outflow data come from Baidu Migration. The control variables at the city level come from China City Statistical Yearbook. In addition, cities without outbreaks were also excluded. After collation, 6417 observations from 301 cities were finally obtained.

## Footnotes

2 Keywords: the novel coronavirus, pneumonia, Zhong Nanshan, symptoms of pneumonia, masks, and correct wearing of masks.

